# Cost-Effectiveness of a Pediatric Operating Room Installation in Sub-Saharan Africa

**DOI:** 10.1101/2023.03.02.23286697

**Authors:** Ava Yap, Salamatu I. Halid, Nancy Ukwu, Ruth Laverde, Paul Park, Greg Klazura, Emma Bryce, Maija Cheung, Elliot Marseille, Doruk Ozgediz, Emmanuel A. Ameh

## Abstract

The unmet need for pediatric surgery incurs enormous health and economic consequences globally, predominantly shouldered by Sub-Saharan Africa (SSA) where children comprise almost half of the population. Lack of economic impact data on improving pediatric surgical infrastructure in SSA precludes informed allocation of limited resources towards the most cost-effective interventions to bolster global surgery for children. We assessed the cost-effectiveness of installing and maintaining a pediatric operating room in a hospital in Nigeria with a pre-existing pediatric surgical service by constructing a decision tree model of pediatric surgical delivery at this facility over a year, comparing scenarios before and after the installation of two dedicated pediatric operating rooms (ORs), which were funded philanthropically. Health outcomes measured in disability-adjusted life years (DALYs) averted were informed by the hospital’s operative registry and prior literature. A societal perspective included costs incurred by the hospital system, charity, and patients’ families. Costs were annualized and reported in 2021 United States dollars ($). The incremental cost-effectiveness ratios (ICERs) of the annualized OR installation were presented from charity and societal perspectives. One-way and probabilistic sensitivity analyses were performed. We found that the installation and maintenance of two pediatric operating rooms averted 1145 DALYs and cost $155,509 annually. Annualized OR installation cost was $87,728 (56% of the overall cost). The ICER of the OR installation was $152 per DALY averted (95% uncertainty interval [UI] 147-156) from the societal perspective, and $77 per DALY averted (95% UI 75-81) from the charity perspective. These ICERs were well under the cost-effectiveness threshold of the country’s half-GDP per capita in 2020 ($1043) and remained cost-effective in one-way and probabilistic sensitivity analyses. Installation of additional pediatric operating rooms in SSA with pre-existing pediatric surgical capacity is therefore very cost-effective, supporting investment in children’s global surgical infrastructure as an economically sound intervention.

## Introduction

The Lancet Commission on Global Surgery strengthened the recognition of surgical care as an essential healthcare intervention in low- and -middle-income countries (LMICs). However, children’s surgery has been dubbed the “unborn, neglected stepchild of global health.”(1) The persistent perception of surgery in LMICs as prohibitively expensive is a leading factor limiting investment in this area. As a result, 1·7 billion children continue to lack access to surgical care worldwide, with 65% or 1·1 billion of them residing in LMIC.(2) This issue particularly affects Sub-Saharan Africa (SSA), where 47% of the population is under 15 years old.(3)

Inadequate pediatric surgical care leads to preventable years of life lost and years lived with disability.(4) A 5-year retrospective review of 1,313 neonatal admissions to a Ugandan hospital estimated 98% unmet needs in treating congenital anomalies.(5) A cohort study spanning 19 SSA countries found that infant mortality from pediatric surgical conditions was markedly higher than that of HICs, especially for gastroschisis (76% vs 20%) and anorectal malformations (11% vs 3%).(6)

Nigeria is the most populous country in SSA, with an estimated population of 211 million and approximately half of whom are aged 18 or younger. Although pediatric surgery is a credentialed subspecialty, the burden of pediatric surgical disease in Nigeria remains largely unmet as this service is only provided by a select few hospitals.(7) A nationwide community survey of 1,883 children identified 81 surgical diagnoses and an estimated 2·9 million children were living with surgically correctable diseases in the country.(8) Therefore, increasing the pediatric surgical capacity in this country is a high priority.

The cost-effectiveness of a wide range of pediatric surgical conditions in LMICs has been previously reported.(9,10) However, this work analyzed specific operations which may be more difficult to apply to strategic health systems development. For example, a cleft repair can be funded widely across LMICs in a vertical approach with substantial economic benefit, but this approach cannot tackle a broad range of diseases encountered by children.(11)

A non-governmental organization (NGO) recognized this gap in capacity and has supported the equipping of pediatric operating rooms (OR) in LMICs to increase surgical infrastructure. This intervention was highly cost-effective in the first OR installation in Uganda, but the results have not been validated or replicated elsewhere.(12) Here we report a CEA of two operating room installations adding to a pre-existing pediatric surgical service in Nigeria.

## Methods and Materials

### Study Setting and Participants

The study is set in a 450-bed national referral hospital in Nigeria that accepts patients from across the country and in the West SSA region. Facilities include a neonatal unit and pediatric surgery service staffed by three accredited pediatric surgeons. In August 2019, two dedicated pediatric operating rooms were installed philanthropically within the hospital grounds to bolster its pediatric surgical infrastructure and alleviate the surgical backlog. Since then, the pediatric surgical volume and case complexity has significantly increased.(13) This CEA incorporates this increased surgical volume, as additional disease burden was averted after the installation of the pediatric OR.

We utilized a prospective perioperative patient clinical registry as part of the collaboration between the hospital and the NGO, stored in a secure database hosted by REDCap, as reported previously.(13,14) Participants were children under the age of 18 who underwent surgery performed by the hospital’s pediatric surgical service from June 2018 to September 2021. Institutional Review Board approvals were obtained from involved institutions for this portion of the study.

### Ethics Statement

Abuja National Hospital (NHA/EC/071/2019) and University of California San Francisco (19-29663) Institutional Review Board approvals were obtained for the purposes of this study. Formal verbal consents were obtained from the parent or guardian of the child beforehand.

### Model and Time Horizon

A decision tree model was constructed in TreeAge Software Version 19.0 to map out the life trajectories of pediatric surgical patients with or without surgical treatment, based on the patient care delivered in the hospital. Model simulations were carried out with Visual Basic for Application (VBA, Version 7·1·1119). In this model, a single decision node (D1) represented the presence or absence of the additional pediatric ORs. Two scenarios were compared: 1) the baseline surgical capacity without a dedicated pediatric OR and 2) the surgical capacity with the additional dedicated pediatric ORs.(Supplementary Materials, Fig S1) The model encapsulated the cost-effectiveness of the incremental OR facility through one year of service. Therefore, the annual number of cases was included as the outcome metrics, as were annualized costs of OR operation and maintenance. The model follows the requirements of the Consolidated Health Economic Evaluation Reporting Standards 2022.(15) Model assumptions are listed in Table 1.

**Table 1:**
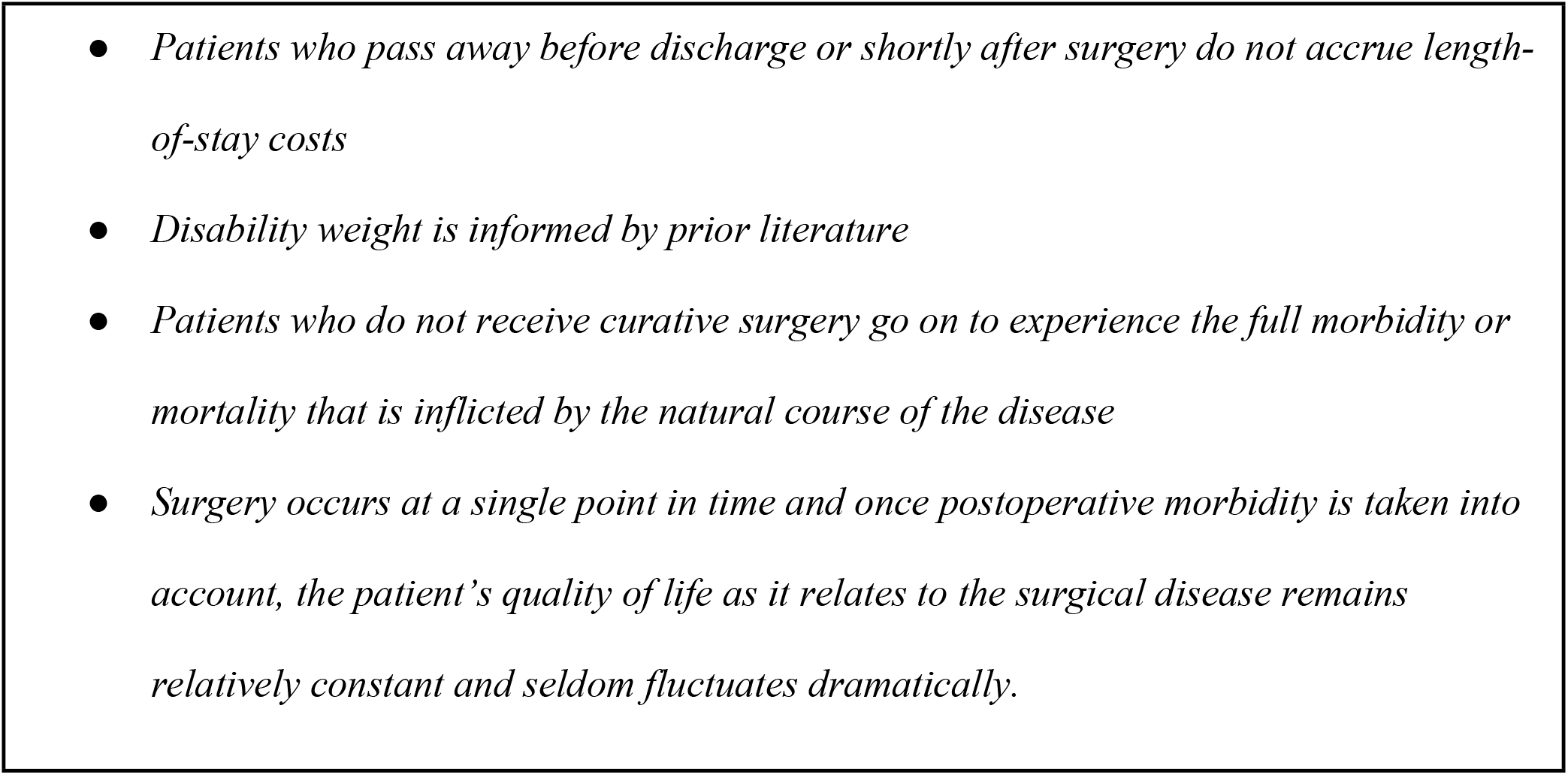
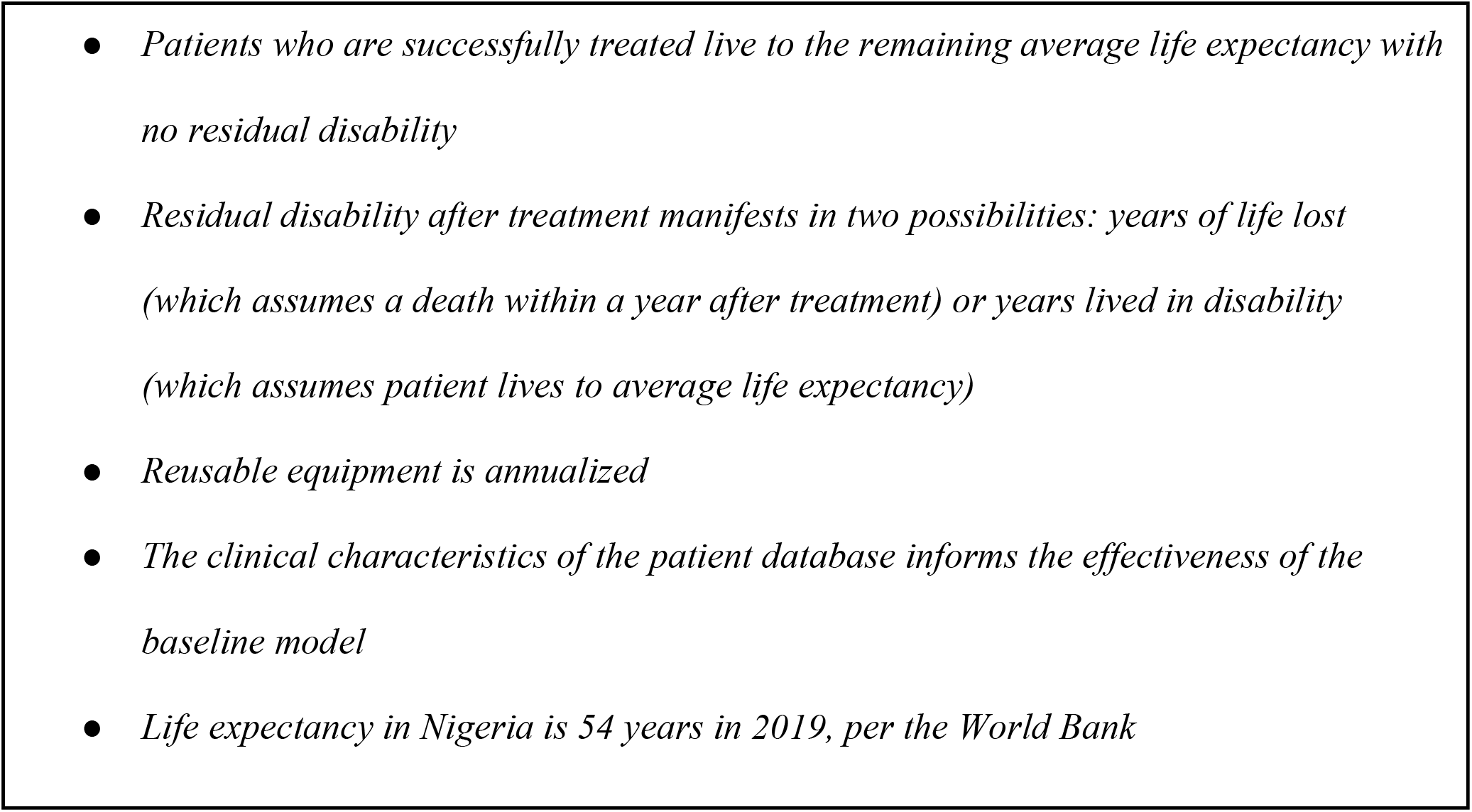
Model assumptions for estimating the cost-effectiveness ratios of a pediatric operating room installation in Nigeria.

### Counterfactual

The counterfactual scenario represented the number and case mix of surgeries performed by the pediatric surgical service during the pre-installation phase. Without a dedicated pediatric OR, only a proportion of patients underwent surgery (66% of the post-installation volume) given insufficient surgical infrastructure. Given the lack of OR infrastructure, patients who could not undergo surgery (nearly 34%) therefore suffered the natural course of the disease as no alternative medical therapy exists.

### Effectiveness Inputs

Health effectiveness was informed by the hospital’s perioperative patient REDCap registry, which holds case counts by disease diagnosis at the time of surgery, and post-operative outcomes including death or successful discharge after surgery. We calculated the health effects of surgical treatment using disability-adjusted life years (DALYs) and lives saved. Disease-specific disability weights were informed by previously published literature. (Supplementary Materials, Table S1) In our sensitivity analysis, we subject DALYs to a time discount of 3% and age weighting of 4%, as recommended by Fox-Rushby et al.

### Cost Inputs

The CEA’s healthcare system perspective was an aggregation of multiple fixed and variable costs of a functional pediatric OR. Costs were reported in 2021 United States dollars ($), using purchasing power parity (PPP) for currency exchange.

Fixed costs included reusable equipment funded by the NGO and perioperative staff salaries funded by the hospital. Reusable and large-scale operative equipment costs were annualized over each item’s projected lifespan as informed by product warranties or reported estimated lifetimes.(16,17)(Table S2) Annual salary costs were informed by public salary scales and verified by the perioperative staff employed at the hospital, then weighted by the proportion of each staff member’s presence per case. (Supplementary Materials, Table S5)

Variable case-based costs included perioperative medications, disposable surgical equipment, postoperative inpatient hospital admission, and family out-of-pocket (OOP) expenses. The anesthetic medication and disposable equipment costs were informed by hospital pharmacy and supply price lists (Supplementary Materials, Table S3, S4) Hospital length of stay costs were derived from the country-specific WHO-CHOICE average. OOP costs were obtained from a self-reported caregiver survey recorded in the REDCap registry.

### Cost-Effectiveness Analysis

Our primary metric for cost-effectiveness is the incremental cost-effectiveness ratio (ICER), which is defined as the (Cost_OR installation_ – Cost_No pediatric OR_)/(DALYs_OR installation_ – DALYs_No pediatric OR_), reported in $ per DALY averted. Per the most recent guidelines of cost-effectiveness threshold, our intervention was deemed cost-effective if the ICER was lower than half the country’s GDP, which is the threshold of choice for resource-constrained areas and suggested by the Disease Control Priorities 3^rd^ edition.(18,19) In Nigeria, this cut-off at half-GDP per capita was $1,043 in 2020 based on the most recent World Bank Data. To put our findings into the current context, this ICER was compared to other ICERs of similar and commonly funded public health interventions.

One-way sensitivity analysis was performed by individually adjusting relevant variables with inherent uncertainty over a plausible range to evaluate how much the ICER changes with each scenario. The societal ICER was used as the reference point. Results were presented in a tornado diagram.

Probabilistic sensitivity analysis was conducted using 100 Monte Carlo simulation batches of yearly caseloads. Parameters with inherent uncertainty were randomized using continuous probability distributions.(Table 2) Ranges are informed by prior studies or +/-0·2 when unavailable, based on the previous methodology.(12,20) ICER uncertainty intervals were obtained by bootstrapping over 100 samples. Results from the probabilistic sensitivity analysis were depicted in cost-effectiveness planes.

**Table 2:** Parameters with inherent uncertainty and their respective probability distributions used for the Monte Carlo simulation used in the probabilistic sensitivity analysis.

| Parameters | Category | Distribution | Source |
| --- | --- | --- | --- |
| Number of annual cases | Effectiveness | Uniform | Hospital database |
| Disability weights | Effectiveness | Beta | Various (Table S1) |
| Probability of death before discharge | Effectiveness | Beta | Hospital database |
| Probability of successful treatment | Effectiveness | Beta | Operative log |
| Long-term equipment costs | Cost - Fixed | Gamma | Charity price sheets |
| Shipping and installation cost | Cost - Fixed | Gamma | Charity price sheets |
| Personnel salaries | Cost - Fixed | Gamma | Public salary scales |
| Disposable equipment cost | Cost- Variable | Gamma | Hospital price sheets |
| Perioperative medication cost | Cost- Variable | Likelihood | Hospital price sheets |
| Inpatient hospital admission cost | Cost- Variable | Gamma | WHO-CHOICE tool |
| Out-of-pocket patient cost | Cost- Variable | Gamma | Hospital database |

## Results

Over 3.2 years, the pediatric surgical surgery service at the hospital performed a total of 1,068 procedures, or a mean of 334 cases per year. 246 (23%) cases were performed before installation of the additional pediatric ORs (June 2018-Aug 2019), whereas 822 (77%) cases were performed post-installation (Aug 2019 - Sept 2021). Of all the cases performed, 806 (75.5%) were elective and 242 (23%) were emergencies. General pediatric surgical disease comprised most diagnoses (689, 65%), distantly followed by congenital anomalies (147, 14%).

From the charity perspective, the annualized cost of donated reusable equipment for two ORs was £37,953 in 2019 uninflated British pounds or $57,435, while the cost of shipping and installation was £19,500 in 2019 uninflated British pounds or $30,293. In the base case analysis, the incremental cost of personnel annual salaries was $22,062. The annual sum of variable costs was $64,324, which included costs of disposable equipment, perioperative medications, inpatient hospital admission, and OOP expenses. The annualized shipping and installation cost for the pediatric ORs was $30,294. The total annualized cost of installation and maintenance of the pediatric operating rooms was $174,114 from the societal perspective or $ 87,728 from the charity perspective. The charity-funded costs made up 50% of the total cost of the intervention.

From the health outcomes standpoint, installation of the pediatric OR averted 1144 DALYs annually in the base case analysis. This amounted to 481 DALYs averted if a 3% time discount was applied, or 703 DALYs averted if an additional 4% age weight was applied.

In the base case scenario, the OR intervention ICER was $152 per DALY averted or $8,215 per life saved from the societal perspective. From the charity or installation perspective, the ICER was $77 per DALY averted or $4,139 per life saved. Notably, both ICERs were well below the cost-effectiveness threshold of half-GDP per capita in Nigeria.

In the one-way univariate sensitivity analysis, the ICER was most sensitive to the proportion of cases that were performed before relative to after installation, ranging from $94-331 per DALY averted when the range of cases was between 0-90% of surgeries. The ICER was also sensitive to the presence of time discounting ($152-361) and age weighting ($152-247). Conversely, the ICER was relatively insensitive to currency exchange methodology, changes in life expectancy, cost of reusable equipment, cost of hospital stay, and salary.(Fig 1)

**Fig 1:**
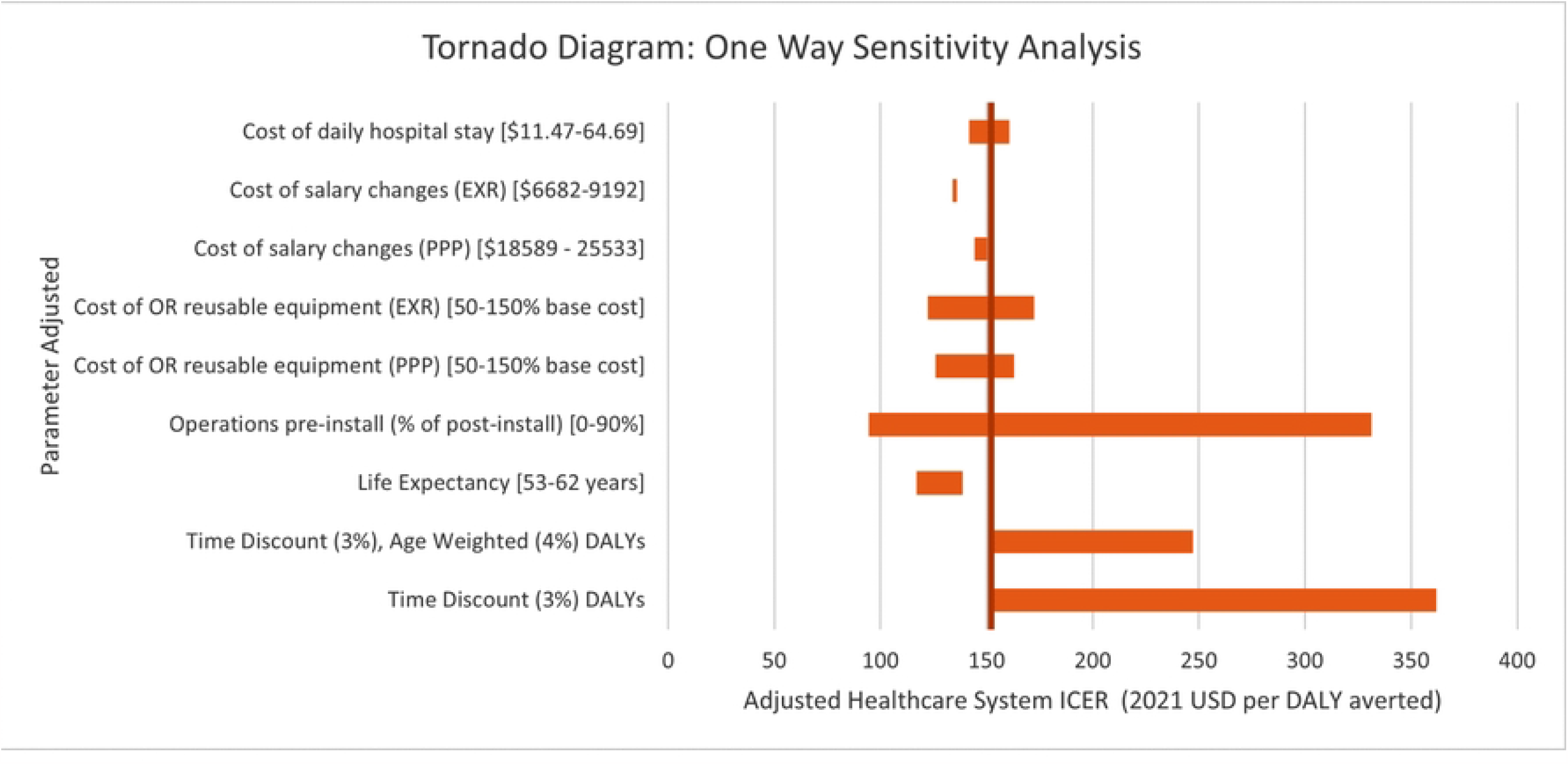
Tornado diagram. Showcases one-way sensitivity analysis of the cost-effectiveness model by adjusting parameters with inherent uncertainty.

In probabilistic sensitivity analysis utilizing Monte Carlo simulations, the pre-installation scenario cost $111,145 (95% confidence interval [CI] 107,968-114,321) and accrued 1,475 DALYs (95% CI 1429-1521). The post-installation scenario cost $282,293 (95% CI 277,440-287,145) and accrued 346 DALYs (95% CI 330-362). The ICER of the OR installation from the societal perspective was $152 per DALY averted (95% uncertainty interval [UI] 147-156). The ICER from the charity perspective was $78 (95% UI 75-81). Both ICERs and respective uncertainty intervals from the charity and societal perspective remained robust and cost-effective.(Fig 2)

**Fig 2:** Cost-effectiveness planes. Planes show the incremental changes in cost and disability-adjusted life years averted. The solid grey line reflects the incremental cost-effectiveness ratio (ICER), while the dotted grey line is the cost-effectiveness threshold at half of the country’s gross domestic product (GDP). The ICER is displayed from the charity perspective in 4a and from the societal perspective in 4b. Both ICERs are well under the half-GDP cost-effectiveness threshold.

**Fig 2A:**
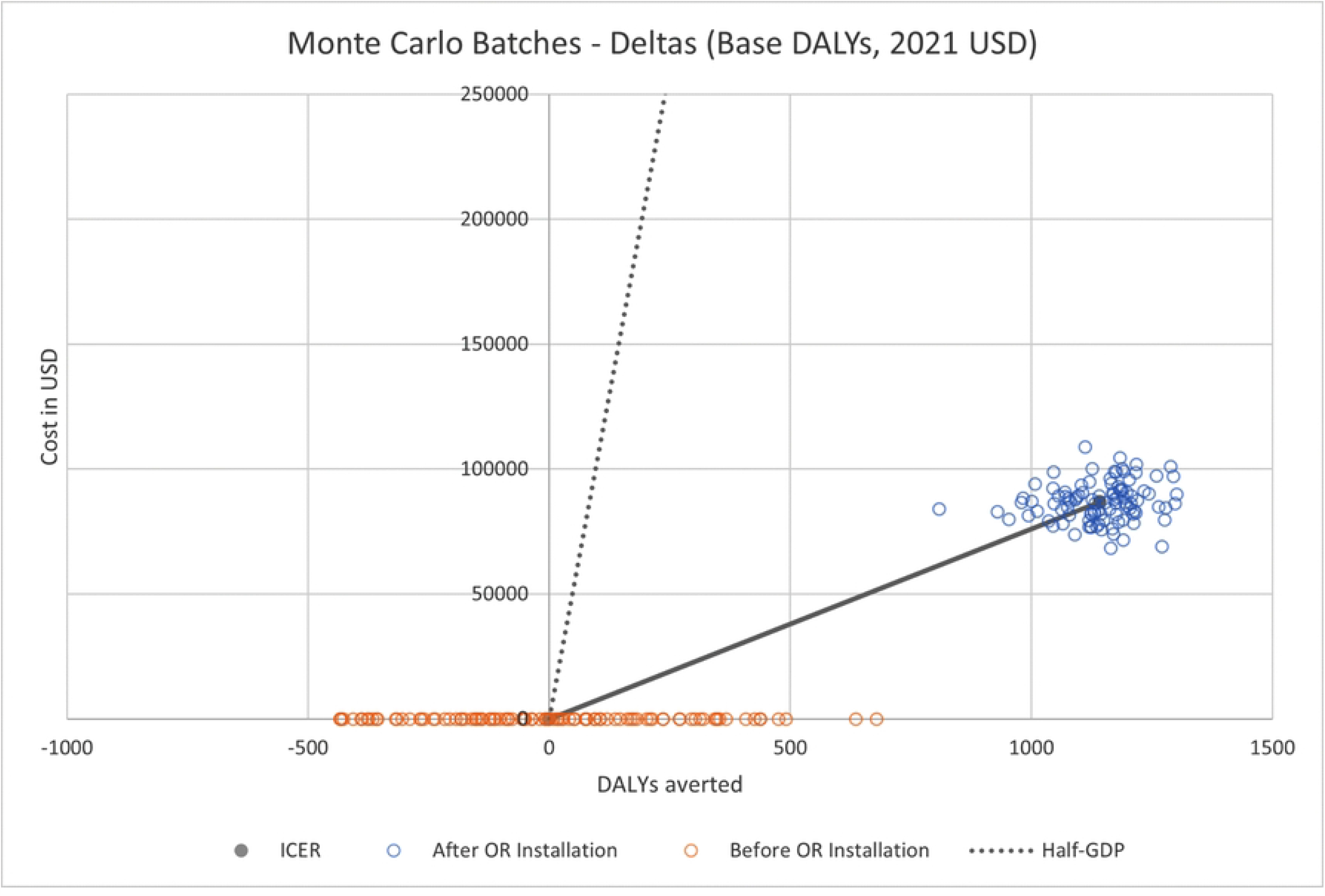
cost-effectiveness plane - charity perspective.

**Fig 2B:**
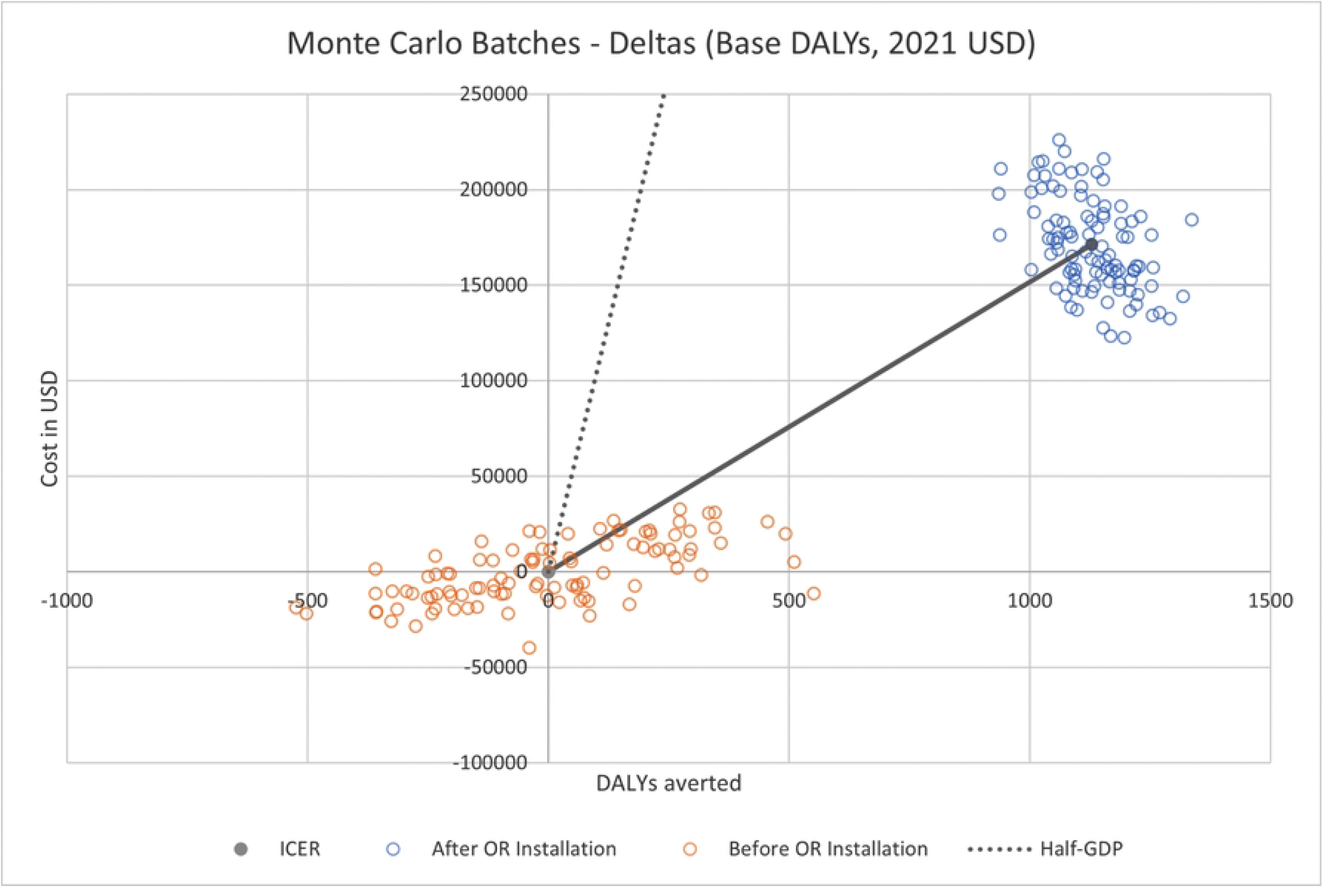
cost-effectiveness plane - societal perspective.

## Discussion

We found that compared to no installation, a dedicated pediatric OR installation in an LMIC hospital with a pre-existing pediatric surgical service is very cost-effective. Utilizing retrospective and prospective data collection, we incorporated real individual clinical outcomes, allowing for a more accurate representation of the disease burden averted. The multidimensional sensitivity analysis reaffirmed the robustness of the model, instilling confidence in the ICER estimates.

Both ICERs from the charity perspective ($77 per DALY averted) and societal perspective ($152 per DALY averted) were lower than half of the country’s GDP-per-capita cutoff ($1043 in 2020), which is a cost-effectiveness threshold proposed for LMICs(18,21) This study’s ICERs were also compared to that of prior cost-effectiveness studies involving pediatric surgeries or other public health initiatives.(Fig 3) The OR installation was at least as cost-effective as other children’s surgery interventions, and more cost-effective than cesarean sections, medical therapy for cardiac conditions, and antiretroviral therapy for human immunodeficiency virus (HIV).(10,22) This finding is pertinent for global health, as it showcases the relative economic efficiency of the OR installation to other well-established interventions that have received a disproportionate share of global health funding.

**Fig 3:**
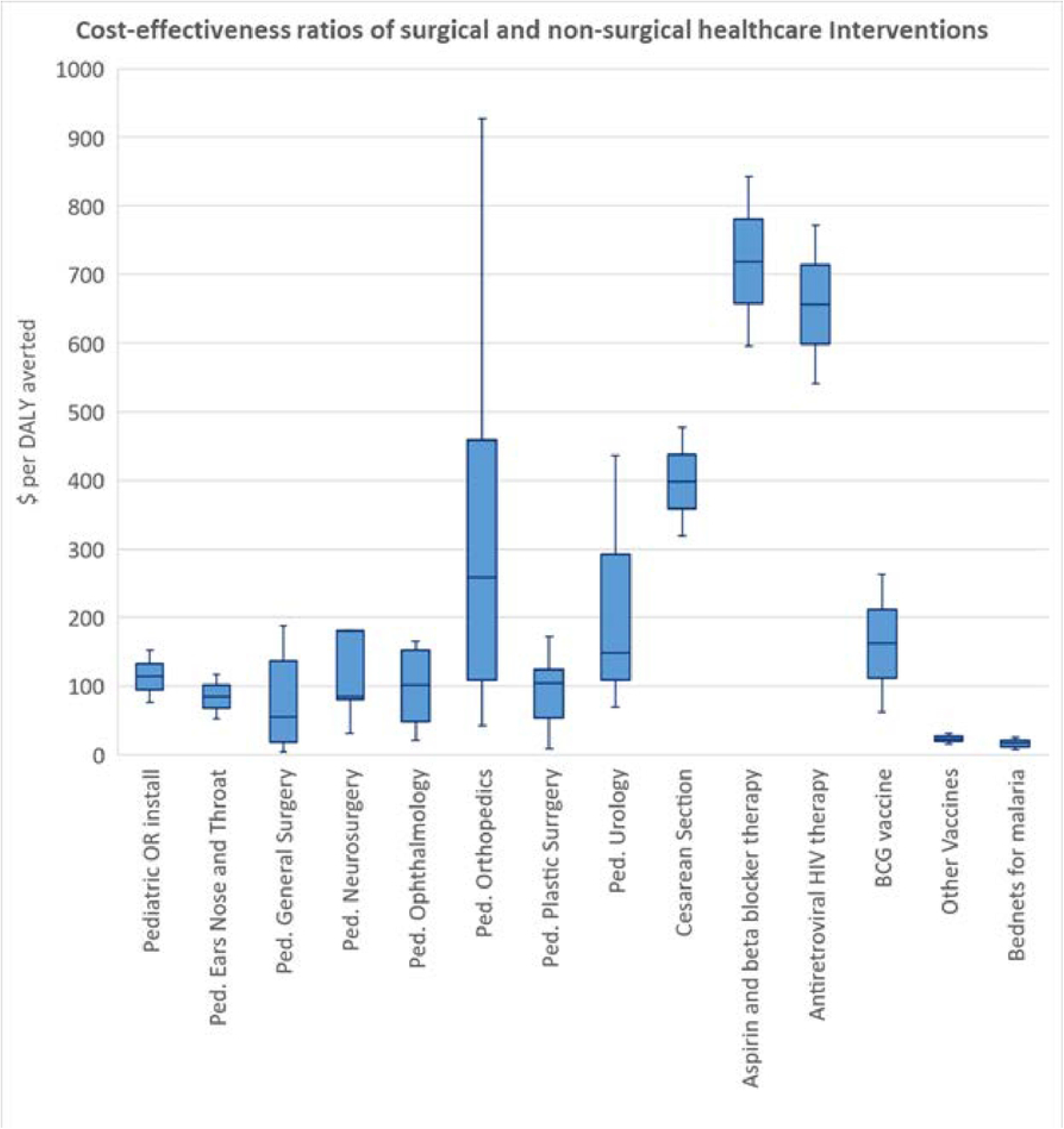
Comparison of the incremental cost-effectiveness ratios (ICERs) across multiple surgical, medical, and public health interventions, including subspecialty pediatric (Ped.) surgeries. Pediatric operating room (OR) installation remains one of the more cost-effective strategies.

Notably, preexisting cost-effectiveness studies in LMICs were almost always conducted for disease-specific interventions. For example, our ICER’s comparison to otolaryngology interventions was predominantly cleft lip or palate repairs. Most disease-specific studies omitted costs of reusable equipment, installation, and patient out-of-pocket costs, components that were included in our study and informed by empirical, patient-derived data.(23–25) Other philanthropic initiatives such as mission trips are less likely to capture such a diversity of costs when focusing solely on charity operational expenses.(26) By including multiple stakeholders, all of whom were necessary to operate the pediatric OR, our CEA provided a more comprehensive and conservative estimate, as we were able to accommodate multiple stakeholders. This societal perspective reflects the reality of a charity-sponsored public health intervention, recognizing that OR installation does not occur in a vacuum and that local surgical support is necessary.

This study’s cost-effectiveness is higher than that of another CEA we conducted for an OR installation in Uganda, at $37·35 per DALY averted.(12) This difference can be attributed to several site characteristics. First, Nigeria’s lower life expectancy (54 years) compared to Uganda’s (63 years) led to a lower number of years saved per surgery. Second, the comparator (counterfactual) scenario in Nigeria was different from that of Uganda. Nigeria had a pre-existing, albeit reduced pediatric surgical volume before the OR installation, while Uganda’s model did not have any surgical volume before installation, so all surgeries performed after OR installation were considered incremental health benefits, leading to more DALYs averted. Third, the pediatric surgery service in the Nigerian hospital saw a larger portion of elective surgeries, with fewer DALYs averted per case. These distinctions demonstrate the heterogeneity in the economic and health impact of an infrastructure-building project in different healthcare systems and countries, substantiating the need to conduct CEA evaluation in a variety of settings to validate its economic viability. In both cases, the pediatric OR installation CEA scenarios mirrored the site-specific situation, so that their ICERs remain true to the respective sites.

Beyond the economic effects of this OR installation, other benefits have been observed within the vicinity of the Nigerian hospital. Enhanced capacity meant time delays in surgery were enormously reduced, effectively eliminating wait times that previously extended to years. This was also true for emergency surgeries, where protracted wait times led to more morbidity. Additionally, since pediatric operating instruments were provided as part of the installation package, surgeries could be conducted with reduced operating times given ready access to the appropriate tools. The consequences of a long surgical waitlist can range from frustration to fatalities.(1,27) More complex surgeries were also conducted on younger infants with more medical comorbidities and higher ASA class with no noticeable change in mortality.(13) Together, these improvements to patient care have augmented perioperative staff morale.

While these are empirical observations noted by the surgical team, a qualitative study involving focus groups and in-depth interviews of the patients’ families and staff, after OR installation can better elucidate and expound on these positive effects, and these efforts are currently ongoing.

Before this OR installation, no dedicated pediatric OR existed in Nigeria, forcing children to compete with the adult population to receive surgery in limited adult ORs lacking the appropriate pediatric instrumentation and exacerbating the unmet need of children’s surgical conditions. This is a common scenario in many LMIC hospitals that lack dedicated ORs for children. As a result of insufficient and delayed care, LMIC pediatric patients fare substantially worse after surgery than those in HIC. For example, surgical congenital anomalies have mortality rates as high as 80% in LMICs and many times higher than that of HIC, which is consistently under 10%.(6,28,29) In Nigeria, neonatal surgical mortality reaches 26·2%, with mortality from gastroschisis at 58·3%, esophageal atresia at 56·5%, and intestinal atresia at 37·2%, based on a recent prospective cohort study of 17 tertiary hospitals.(30)

Nigeria is one of few LMICs that have published a national surgical, obstetric, anesthesia, and nursing plan after the World Health Assembly mandated that countries provide essential surgical care and anesthesia as part of their universal health coverage package.(31,32) However, committed investment to bolster pediatric surgical services are still lacking, as funding for pediatric surgery in SSA continues to stall.(33) Installing a pediatric operating room is only one way to increase the surgical capacity, but true progress can only be achieved through a multilateral public health initiative that encompasses infrastructure improvement, workforce expansion, and financial coverage for patients’ healthcare expenses. NGOs have provided charity-sponsored surgeries for patients with cleft deformities and pediatric surgical training stipends.(34,35) However, to ensure sustainability in these practices, local governments and Ministries of Health will also need to take steps toward investing in this area.

Limitations to the study include the theoretical construct of the decision tree model. Its effectiveness rests on the reliance on the proportional increase in cases after operating room installation. Of note, the increase in patient clinical complexity after OR installation was not included in this study, as our analysis only considered a difference in case volume. However, this suggests our estimate of the cost-effectiveness is more conservative, as we did not incorporate surgeries of more complex, disabling diseases that may lead to more DALYs averted. Other assumptions were made in constructing this model, such as the patient’s postoperative course after leaving the hospital, which remained in a steady state to be consistent with the decision tree model. However, since the average life expectancy of Nigeria was used as a benchmark for DALYs averted, this assumption should not skew the results. Finally, although installation costs and initial maintenance labor were accounted for, the analysis excluded charity overhead expenses such as administrative costs as these were difficult to attribute towards a single OR installation project.

## Conclusion

This is the first cost-effectiveness analysis of a pediatric OR installation in Nigeria, Africa’s most populous country. Installation of two dedicated pediatric ORs is deemed very cost-effective with an ICER of $151 per DALY averted from the societal perspective. This ICER is lower and more cost-effective than other essential interventions such as cesarean sections, HIV antiretroviral therapy, and medical therapy for cardiac disease. This inaugural study will open more opportunities for cost-effectiveness analysis and other forms of economic evaluation of pediatric surgical initiatives in Nigeria and other LMICs, as little current research has been done. The findings of this study can serve as a strong advocacy tool for policymakers and funders alike to scale up essential children’s surgical care.

## Data Availability

The data underlying this article are available in the article and in its online supplementary material.

